# Serum gamma-glutamyl transferase level is associated with the risk of pancreatic cystic neoplasms: A nationwide cohort study

**DOI:** 10.1101/2025.03.11.25323676

**Authors:** Min Woo Lee, Jin Myung Park, In Rae Cho, Kwang Hyun Chung, Jin Ho Choi, Woo Hyun Paik, Ji Kon Ryu, Bong Seoung Kim, Kyung Do Han, Sang Hyub Lee

## Abstract

•

**Background/Aims:** Gamma-glutamyl transferase (GGT) is a known surrogate marker of hepatic dysfunction and oxidative stress. However, data on its association with pancreatic disease, especially pancreatic cystic neoplasm, is unknown. This study aimed to investigate the association of GGT with the incidence of pancreatic cystic neoplasm.

**Methods:** Participants who received general health checkup by National Health Insurance Service in 2009 were included. Newly diagnosed cases of pancreatic cystic neoplasms from one year after the health checkup to 2020, the end of the study period, were identified. Participants were divided into quartile groups based on GGT levels. Multivariable cox proportional hazard models were used to estimate the risk of pancreatic cystic neoplasms according to GGT quartile (Q1-Q4).

**Results:** There were 28,940 cases of pancreatic cystic neoplasms among 2,655,665 eligible participants. The incidence rate was 1.09 cases per 1,000 person-years, with a median follow-up of 10.32 (IQR: 10.09-10.58) years. In multivariate regression analysis, adjusted hazard ratios for GGT quartiles using Q1 group as a reference were: 1.043 (95% CI: 1.009-1.079) for Q2, 1.075 (95% CI: 1.039-1.111) for Q3, and 1.138 (95% CI: 1.099-1.178) for Q4.

**Conclusions:** Higher GGT level was associated with increased risk of pancreatic cystic neoplasms. Therefore, serum GGT levels might have a role as a biomarker for the development of PCN.

## INTRODUCTION

Pancreatic cystic neoplasm (PCN) is a heterogeneous group of cystic tumors that include serous cystic adenomas, mucinous cystic neoplasms, intraductal papillary mucinous neoplasms, and rarely solid pseudopapillary tumors [1]. Among them, mucinous cysts have some risk of progressing to invasive carcinoma or developing concomitant pancreatic ductal adenocarcinoma [2, 3, 4, 5]. Therefore, risk stratification and surveillance strategy are always needed for PCN [6, 7].

Progression of PCN to invasive carcinoma has been extensively investigated, including genetic alterations, metabolites, and serologic markers such as CA19-9 [8, 9, 10]. However, few studies have reported the development of PCN [11, 12]. In addition, many guidelines focus on risk stratification and surveillance rather than early detection [6, 13].

With recent improvement in the resolution of cross-sectional imaging, the detection of pancreatic cystic neoplasms has increased over the years [14]. PCNs are usually asymptomatic, and most lesions are identified incidentally on cross-sectional imaging [15]. Many researchers have investigated factors associated with PCN and found that their prevalence is associated with age, BMI, and diabetes [14, 16, 17]. However, it is not known which factors are associated with the incidence of PCN [17].

Gamma-glutamyl transferase (GGT) reflects hepatic dysfunction or oxidative stress. It has recently been reported to be associated with metabolic diseases, cardiovascular diseases, and malignancies [18, 19, 20]. Furthermore, some researchers have found an association between GGT and pancreatic cancer [21, 22]. However, data on the association between GGT and PCN are still limited. Therefore, this study aimed to investigate the association between GGT and incidence of PCN in a nationwide population-based cohort with long-term surveillance.

## METHODS

### Data collection

This nationwide retrospective cohort study utilized data from 2.8 million people who underwent general health checkups by the National Health Insurance Service (NHIS) of South Korea in 2009. The NHIS is the national healthcare system that covers more than 97% of South Koreans. The NHIS database was compiled for the purpose of reimbursing healthcare providers. It consists of a wide range of health information such as demographics, socioeconomic status, diagnoses, surgeries, prescriptions, and health checkups [23, 24]. In this study, diagnosis histories for pancreatitis, pseudocysts, pancreatic cancer, and pancreatic cystic neoplasms were extracted from the National Health Insurance Service database using the International Classification of Diseases–10th Revision, Clinical Modification (ICD-10-CM) diagnostic codes (Table S1).

The NHIS has been providing free health checkups biannually after the age of 40 since 2009. This health checkup includes anthropometric measurements (height, weight, waist circumference), blood pressure measurements, laboratory tests, health-related behavioral surveys (smoking, drinking, regular exercise), and medical and family history [25]. Therefore, this study utilized data from the NHIS health checkup and subsequent ten years. This study was approved by the Institutional Review Board (IRB) of Seoul National University Hospital (IRB No. H-2406-052-1542), and reported in accordance with the STROBE guideline [26].

### Study population

The timeline of this study and the process of identifying the cohort are shown in Figure 1. We identified 2,896,383 people aged 40 years or more who had a health checkup between January 1 and December 31, 2009. To exclude individuals who already had PCN, cases diagnosed with pancreatitis/pseudocysts (n = 64,763) or pancreatic cystic neoplasms (n = 4,298) until the date of health checkup were excluded. Participants who had a general health checkup but had missing values for any of the variables used in the study (n = 160,987) were also excluded. Finally, cases diagnosed with PCN or death within a 1-year lag period of enrollment (n = 10,670) were excluded to eliminate the possibility of pre-existing PCN.

**Figure 1.**
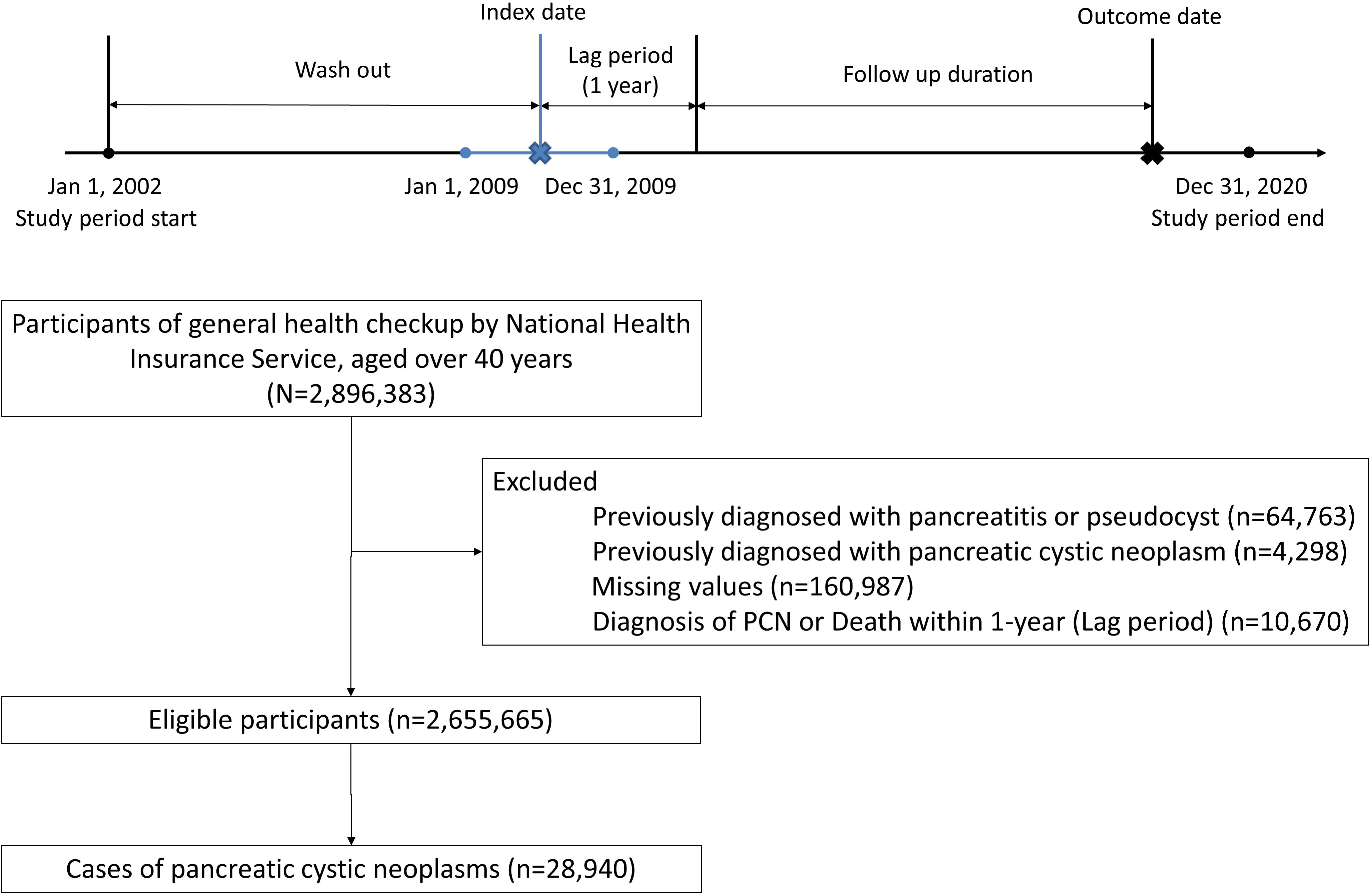
Study population.

### Covariates measurements

The following variables were measured by the clinician during a health checkup: age, sex, height, weight, waist circumference, and systolic/diastolic blood pressure. Body mass index (BMI) was calculated by dividing weight by the square of height (kg/m^2^). Laboratory tests included fasting blood glucose, estimated GFR, liver function tests (AST, ALT, GGT), and lipid profile (total cholesterol, TG, HDL-C, LDL-C). Comorbidities such as hypertension, type 2 diabetes, chronic kidney disease, and dyslipidemia were defined based on ICD-10-CM diagnostic codes, NHIS codes, prescribed medications, and health checkup data (Table S2).

Information on lifestyle variables was collected through standardized questionnaires during health checkups, including smoking history (nonsmoker, ex-smoker, or current smoker), alcohol consumption (none, mild-to-moderate consumption [< 20 g/day for females, < 30 g/day for males], or heavy consumption [≥ 20 g/day for females, ≥ 30 g/day for males]), and regular exercise (intense 20-minute workouts ≥ 3 days weekly or moderate 30-minute workouts ≥ 5 days weekly). Low-income group was defined as being the lowest quartile of income-proportional insurance premiums or receiving medical aid program.

### Study outcomes

The primary outcome was the incidence of newly developed PCNs after index health checkups. PCNs were identified using ICD-10-CM codes and diagnosis dates from the NHIS database (K86.2 for pancreas cyst; D13.6 for cystic neoplasm of pancreas, benign; D37.7 for cystic neoplasm of pancreas, uncertain behavior). Incidence rates of PCN were stratified by sex-specific quartile of serum GGT level (Q1, < 23 IU/L for males and < 14 IU/L for females; Q2, < 34 IU/L for males and < 18 IU/L for females; Q3, < 57 IU/L for males and < 26 IU/L for females; Q4, ≥ 57 IU/L for males and ≥ 26 IU/L for females). The association between GGT level and risk of PCN was evaluated by regression analysis. Subgroup analyses were performed according to age, sex, obesity, lifestyle factors (smoking, alcohol consumption, regular exercise), and comorbidities (hypertension, diabetes mellitus, dyslipidemia, chronic kidney disease).

### Statistical analysis

For continuous variables, normality was assessed using the Shapiro-Wilk test. Continuous variables with normal distribution were presented with mean and standard deviation. Continuous variables without normal distributions were presented with geometric means and 95% confidence intervals. Differences between GGT quartile groups were evaluated using one-way analysis on variance for continuous variables and chi-square tests for categorical variables. Incidence rates for PCN of each quartile were calculated by dividing the number of events by 1000 person-years. The cumulative incidence probability for each quartile group was plotted using Kaplan-Meier curves and compared using the log-rank test. Multivariable Cox proportional regression analyses were used to estimate hazard ratios (HRs) and 95% confidence intervals (CIs) for the association between serum GGT level and the risk of PCN. Model 1 was unadjusted. Model 2 was adjusted for age and sex. Model 3 was adjusted for age, sex, lifestyle factors (smoking, alcohol consumption, regular exercise, income quartile), and comorbidities (diabetes, hypertension, dyslipidemia, chronic kidney disease). A two-sided *P*-value of less than 0.05 was considered statistically significant. Statistical analyses were performed using SAS version 9.3 (SAS Institute Inc., Cary, NC, USA) and R version 4.4.1 (The R Foundation for Statistical Computing, Vienna, Austria).

## RESULTS

### Baseline characteristics

Table 1 presents baseline characteristics of the cohort by quartile of serum GGT level. There were 2,655,665 participants aged 54.3 ± 10.5 years. Regarding the percentage of participants in each quartile, it was 25.5% in Q1, 23.8% in Q2, 25.7% in Q3, and 25.0% in Q4. Those in higher quartiles had higher blood pressure, higher weights, higher waist circumferences, higher BMI, higher rates of current smoking and higher rates of heavy alcohol consumption but lower rates of regular exercise. Higher quartile groups showed higher prevalence of diabetes mellitus, hypertension, and dyslipidemia. In laboratory tests during the NHIS health checkup, serum total cholesterol, triglycerides, and liver enzyme levels were higher in higher quartile groups.

**Table 1.** Baseline characteristics grouped by GGT level.

|  | Serum GGT level |  |  |  | *P*-value |
| --- | --- | --- | --- | --- | --- |
|  | Q1 (N=678,027) | Q2 (N=631,006) | Q3 (N=683,678) | Q4 (N=662,954) |  |
| Age, years | 53.99±11.04 | 54.34±10.57 | 54.54±10.3 | 54.3±9.92 | < 0.0001 |
| Sex (male) | 330522 (48.75) | 326287 (51.71) | 335805 (49.12) | 336263 (50.72) | < 0.0001 |
| Height, cm | 161.72±8.57 | 161.94±8.75 | 161.46±8.94 | 161.51±9.02 | < 0.0001 |
| Weight, kg | 60.08±9.44 | 62.32±10.26 | 63.76±10.8 | 65.23±11.11 | < 0.0001 |
| BMI, kg/m<sup>2</sup> | 22.91±2.68 | 23.68±2.85 | 24.37±3.01 | 24.93±3.2 | < 0.0001 |
| Waist Circumference, cm | 78.19±7.96 | 80.51±8.32 | 82.23±8.47 | 83.94±8.51 | < 0.0001 |
| Systolic BP, mmHg | 120.82±15.02 | 123.24±15.11 | 125.23±15.35 | 127.69±15.79 | < 0.0001 |
| Diastolic BP, mmHg | 74.9±9.87 | 76.57±9.95 | 77.85±10.12 | 79.49±10.45 | < 0.0001 |
| Income, lowest quartile | 139230(20.53) | 126443(20.04) | 137994(20.18) | 135313(20.41) | < 0.0001 |
| Smoking |  |  |  |  | < 0.0001 |
| Non | 466617(68.82) | 404113(64.04) | 431045(63.05) | 388541(58.61) |  |
| Ex | 104231(15.37) | 104399(16.54) | 105572(15.44) | 97486(14.7) |  |
| Current | 107179(15.81) | 122494(19.41) | 147061(21.51) | 176927(26.69) |  |
| Alcohol consumption |  |  |  |  | < 0.0001 |
| Non | 469136(69.19) | 386982(61.33) | 385451(56.38) | 315205(47.55) |  |
| Mild | 192284(28.36) | 214338(33.97) | 244884(35.82) | 243625(36.75) |  |
| Heavy | 16607(2.45) | 29686(4.7) | 53343(7.8) | 104124(15.71) |  |
| Regular exercise | 141972(20.94) | 129964(20.6) | 135520(19.82) | 123807(18.68) | < 0.0001 |
| Diabetes mellitus | 47748(7.04) | 57860(9.17) | 82847(12.12) | 116527(17.58) | < 0.0001 |
| Hypertension | 159803(23.57) | 187639(29.74) | 245357(35.89) | 284908(42.98) | < 0.0001 |
| Dyslipidemia | 85023(12.54) | 117879(18.68) | 168725(24.68) | 204262(30.81) | < 0.0001 |
| Chronic kidney disease | 50370(7.43) | 50632(8.02) | 59473(8.7) | 57527(8.68) | < 0.0001 |
| Fasting glucose, mg/dL | 95.22±19.82 | 97.76±22.77 | 100.74±25.96 | 106.23±31.61 | < 0.0001 |
| Estimated GFR, mL/min/1.73m<sup>2</sup> | 85.84±36.4 | 84.49±38.14 | 84.33±39.19 | 84.99±37.51 | < 0.0001 |
| Total Cholesterol, mg/dL | 189.41±33.53 | 197.61±35.16 | 202.83±37.04 | 206.82±40.37 | < 0.0001 |
| HDL -C, mg/dL | 56.11±29.52 | 55.6±29.27 | 55.45±30.01 | 56.03±29.36 | < 0.0001 |
| LDL -C, mg/dL | 113.67±35.48 | 118.38±37.06 | 119.97±39.03 | 116.79±42.7 | < 0.0001 |
| *Triglyceride, mg/dL | 93.37<br>(93.26-93.48) | 109.43<br>(109.29-109.57) | 125.29<br>(125.13-125.45) | 150.01<br>(149.81-150.22) | < 0.0001 |
| *AST, IU/L | 20.97 | 22.31 | 24 | 29.95 | < 0.0001 |
|  | (20.95-20.98) | (22.29-22.33) | (23.98-24.02) | (29.92-29.98) |  |
| *ALT, IU/L | 16.51<br>(16.5-16.53) | 19.25<br>(19.23-19.27) | 22.47<br>(22.44-22.49) | 30.91<br>(30.87-30.95) | < 0.0001 |

Variables are presented as mean ± standard deviation, median (interquartile range), or number (%). Variables marked with \* are presented with geometric mean and 95% CI.
GGT, gamma-glutamyl transferase; BMI, body mass index; BP, blood pressure; HDL-C, high-density lipoprotein cholesterol; LDL-C, low-density lipoprotein cholesterol; AST, aspartate aminotransferase; ALT, alanine aminotransferase.

### Risk of PCN by serum GGT quartile

During a median follow-up of 10.3 years (IQR: 10.1-10.6 years), 28,940 PCN diagnoses were reported among 2,655,665 participants (Table 2). The incidence rate of PCN was 1.09 per 1000 person-years in the entire cohort. Compared with the Q1 group, higher GGT quartiles were associated with a significantly higher risk of PCN after adjusting for confounders (Q2: adjusted HR [aHR], 1.043 [95% CI, 1.009 to 1.079]; Q3: aHR 1.075 [95% CI, 1.039 to 1.111]; Q4: aHR, 1.138 [95% CI, 1.099 to 1.178]). The incidence probability of PCN was the highest in the Q4 group and the lowest in the Q1 group. This order was maintained throughout the observation period (log-rank *P* < 0.001) (Figure 2).

**Figure 2.**
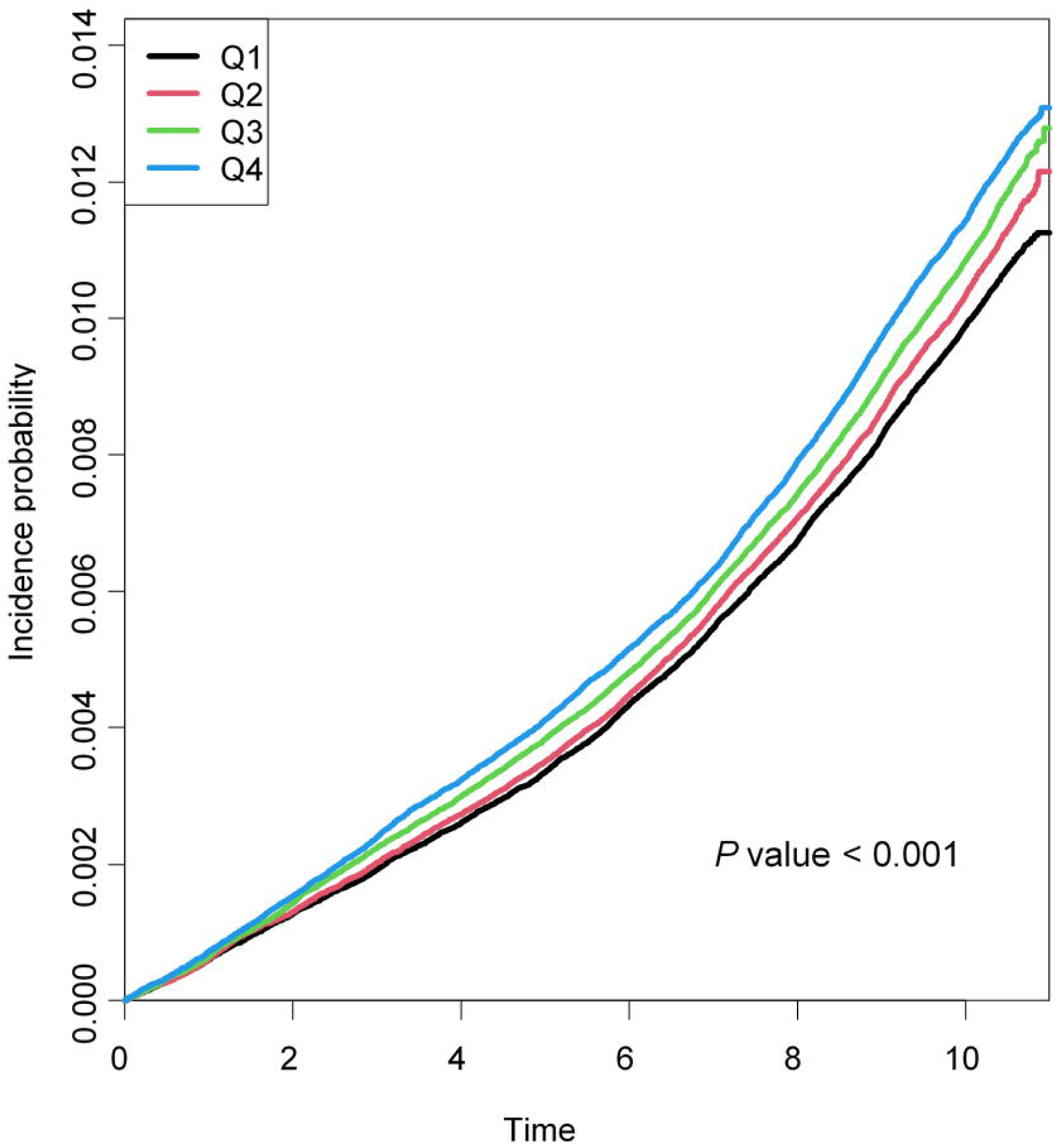
Incidence probability of pancreatic cystic neoplasm stratified by gamma-glutamyl transferase (GGT) level.

**Table 2.** Multivariate regression analysis of the risk of pancreatic cystic neoplasm according to quartile.

| GGT | N | PCN | Duration,<br>years | IR,<br>per 1,000 PY | HR (95% CI) |  |  |
| --- | --- | --- | --- | --- | --- | --- | --- |
|  |  |  |  |  | Model 1 | Model 2 | Model 3 |
| Q1 | 678,027 | 6,864 | 6792685.48 | 1.01 | 1(Ref.) | 1(Ref.) | 1(Ref.) |
| Q2 | 631,006 | 6,736 | 6330561.72 | 1.06 | 1.053 (1.018,1.089) | 1.043 (1.009,1.079) | 1.043 (1.009,1.079) |
| Q3 | 683,678 | 7,649 | 6860249.12 | 1.11 | 1.103 (1.068,1.14) | 1.077 (1.043,1.113) | 1.075 (1.039,1.111) |
| Q4 | 662,954 | 7,691 | 6585301.18 | 1.17 | 1.158 (1.121,1.197) | 1.148 (1.111,1.185) | 1.138 (1.099,1.178) |

Model 1, Not adjusted.
Model 2, Adjusted for age and sex.
Model 3, Adjusted for age, sex, income, BMI, smoking, drinking, regular exercise, Diabetes mellitus,
hypertension, dyslipidemia, CKD.
GGT, gamma-glutamyl transferase; PCN, pancreatic cystic neoplasm; IR, incidence rate; HR, hazard ratio

To ensure the reliability of these results, we further analyzed the association between risk of PCN and more subdivided GGT levels. Dose-response relationship was maintained when serum GGT levels were divided into decile groups (Table S3). All *P*-values for trend were less than 0.001.

### Subgroup analysis

We performed stratified analyses according to pre-specified factors after adjusting for confounders (Table 3). The risk of PCN was compared between the highest serum GGT quartile (Q4) and the rest of the group (Q1-Q3). The Q4 group had a significantly higher risk of PCN than Q1-Q3 groups of all other subgroups except for diabetes (aHR: 0.99 [95% CI: 0.931 to 1.053]). Risk difference was the highest in the nonalcoholic subgroup (aHR: 1.123 [95% CI: 1.085 to 1.163]). There were significant interactions between the risk of PCN and pre-specified subgroups such as obesity, alcohol consumption, and diabetes (*P* for interaction: 0.046, 0.015, and < 0.001, respectively).

**Table 3.** Subgroup analysis.

| Subgroup |  | GGT | N | PCN | Duration,<br>years | IR,<br>per 1,000 PY | HR (95% CI) |  |
| --- | --- | --- | --- | --- | --- | --- | --- | --- |
|  |  |  |  |  |  |  | Univariate | Multivariate* |
| Age | 40-64 | Q1-Q3 | 1608646 | 15427 | 16422259.06 | 0.94 | 1 (Ref.) | 1 (Ref.) |
|  |  | Q4 | 548805 | 5752 | 5540769.65 | 1.04 | 1.108 (1.075,1.142) | 1.07 (1.037,1.104) |
|  | ≥65 | Q1-Q3 | 384065 | 5822 | 3561237.26 | 1.63 | 1 (Ref.) | 1 (Ref.) |
|  |  | Q4 | 114149 | 1939 | 1044531.53 | 1.86 | 1.138 (1.081,1.198) | 1.12 (1.063,1.18) |
|  | p for inter |  |  |  |  |  | 0.3785 | 0.135 |
| Sex | Male | Q1-Q3 | 992614 | 9438 | 9844762.8 | 0.96 | 1 (Ref.) | 1 (Ref.) |
|  |  | Q4 | 336263 | 3081 | 3304098.27 | 0.93 | 0.975 (0.936,1.016) | 1.056 (1.012,1.102) |
|  | Female | Q1-Q3 | 1000097 | 11811 | 10138733.52 | 1.16 | 1 (Ref.) | 1 (Ref.) |
|  |  | Q4 | 326691 | 4610 | 3281202.91 | 1.40 | 1.209 (1.168,1.251) | 1.113 (1.074,1.152) |
|  | p for inter |  |  |  |  |  | <.0001 | 0.0586 |
| Obesity | No | Q1-Q3 | 1380869 | 14468 | 13822858.61 | 1.05 | 1 (Ref.) | 1 (Ref.) |
|  |  | Q4 | 346198 | 4046 | 3409592.7 | 1.19 | 1.138 (1.099,1.178) | 1.113 (1.074,1.153) |
|  | Yes | Q1-Q3 | 611842 | 6781 | 6160637.72 | 1.10 | 1 (Ref.) | 1 (Ref.) |
|  |  | Q4 | 316756 | 3645 | 3175708.48 | 1.15 | 1.044 (1.003,1.087) | 1.054 (1.012,1.098) |
|  | p for inter |  |  |  |  |  | 0.0016 | 0.0464 |
| Smoke | Non, Ex | Q1-Q3 | 1615977 | 18150 | 16245614.46 | 1.12 | 1 (Ref.) | 1 (Ref.) |
|  |  | Q4 | 486027 | 6179 | 4848109.02 | 1.27 | 1.143 (1.11,1.176) | 1.089 (1.057,1.122) |
|  | Current | Q1-Q3 | 376734 | 3099 | 3737881.86 | 0.83 | 1 (Ref.) | 1 (Ref.) |
|  |  | Q4 | 176927 | 1512 | 1737192.16 | 0.87 | 1.052 (0.99,1.119) | 1.091 (1.025,1.162) |
|  | p for inter |  |  |  |  |  | 0.0175 | 0.9563 |
| Alcohol<br>consumption | Non | Q1-Q3 | 1241569 | 14499 | 12405702.8 | 1.17 | 1 (Ref.) | 1 (Ref.) |
|  |  | Q4 | 315205 | 4482 | 3128550.13 | 1.43 | 1.227 (1.187,1.269) | 1.123 (1.085,1.163) |
|  | Mild | Q1-Q3 | 651506 | 5861 | 6579284.89 | 0.89 | 1 (Ref.) | 1 (Ref.) |
|  |  | Q4 | 243625 | 2243 | 2430009.54 | 0.92 | 1.04 (0.99,1.092) | 1.03 (0.981,1.082) |
|  | Heavy | Q1-Q3 | 99636 | 889 | 998508.63 | 0.89 | 1 (Ref.) | 1 (Ref.) |
|  |  | Q4 | 104124 | 966 | 1026741.52 | 0.94 | 1.061 (0.969,1.163) | 1.066 (0.973,1.168) |
|  | p for inter |  |  |  |  |  | <.0001 | 0.0148 |
| Regular<br>Exercise | No | Q1-Q3 | 1585255 | 16599 | 15878455.3 | 1.05 | 1 (Ref.) | 1 (Ref.) |
|  |  | Q4 | 539147 | 6182 | 5350376.36 | 1.16 | 1.108 (1.076,1.141) | 1.094 (1.062,1.128) |
|  | Yes | Q1-Q3 | 407456 | 4650 | 4105041.02 | 1.13 | 1 (Ref.) | 1 (Ref.) |
|  |  | Q4 | 123807 | 1509 | 1234924.82 | 1.22 | 1.081 (1.02,1.146) | 1.071 (1.009,1.135) |
|  | p for inter |  |  |  |  |  | 0.4652 | 0.504 |
| Diabetes<br>mellitus | No | Q1-Q3 | 1804256 | 18402 | 18175069.52 | 1.01 | 1 (Ref.) | 1 (Ref.) |
|  |  | Q4 | 546427 | 6069 | 5462111.66 | 1.11 | 1.1 (1.068,1.132) | 1.114 (1.081,1.148) |
|  | Yes | Q1-Q3 | 188455 | 2847 | 1808426.8 | 1.57 | 1 (Ref.) | 1 (Ref.) |
|  |  | Q4 | 116527 | 1622 | 1123189.52 | 1.44 | 0.916 (0.862,0.974) | 0.99 (0.931,1.053) |
|  | p for inter |  |  |  |  |  | <.0001 | 0.0006 |
| Hypertension | No | Q1-Q3 | 1399912 | 13423 | 14178760.31 | 0.95 | 1 (Ref.) | 1 (Ref.) |
|  |  | Q4 | 378046 | 3939 | 3795697.71 | 1.04 | 1.099 (1.06,1.138) | 1.105 (1.066,1.146) |
|  | Yes | Q1-Q3 | 592799 | 7826 | 5804736.01 | 1.35 | 1 (Ref.) | 1 (Ref.) |
|  |  | Q4 | 284908 | 3752 | 2789603.47 | 1.34 | 0.998 (0.96,1.038) | 1.071 (1.03,1.115) |
|  | p for inter |  |  |  |  |  | 0.0003 | 0.2515 |
| Dyslipidemia | No | Q1-Q3 | 1621084 | 16422 | 16279851.44 | 1.01 | 1 (Ref.) | 1 (Ref.) |
|  |  | Q4 | 458692 | 5021 | 4548820.32 | 1.10 | 1.098 (1.063,1.133) | 1.107 (1.071,1.144) |
|  | Yes | Q1-Q3 | 371627 | 4827 | 3703644.88 | 1.30 | 1 (Ref.) | 1 (Ref.) |
|  |  | Q4 | 204262 | 2670 | 2036480.86 | 1.31 | 1.006 (0.96,1.055) | 1.053 (1.003,1.104) |
|  | p for inter |  |  |  |  |  | 0.0027 | 0.0831 |
| CKD | No | Q1-Q3 | 1832236 | 19256 | 18437941.67 | 1.04 | 1 (Ref.) | 1 (Ref.) |
|  |  | Q4 | 605427 | 6939 | 6034294.22 | 1.15 | 1.104 (1.074,1.134) | 1.097 (1.066,1.129) |
|  | Yes | Q1-Q3 | 160475 | 1993 | 1545554.65 | 1.29 | 1 (Ref.) | 1 (Ref.) |
|  |  | Q4 | 57527 | 752 | 551006.96 | 1.36 | 1.06 (0.975,1.153) | 1.026 (0.943,1.116) |
|  | p for inter |  |  |  |  |  | 0.3681 | 0.1369 |

GGT, gamma-glutamyl transferase; PCN, pancreatic cystic neoplasm; IR, incidence rate; HR, hazard ratio

## DISCUSSION

In this nationwide cohort study of more than 2 million participants, it was found that higher serum GGT levels were associated with an increased risk of PCN. The dose-response relationship was identified in both quartile and decile groups. In subgroup analyses, the association between serum GGT level and PCN risk was the most pronounced in the non-alcoholic group.

This nationwide cohort study is the first to provide evidence that higher serum GGT levels are associated with PCN risk in the general population. We further categorized serum GGT levels, including those within the normal range and found dose-response relationships. The incidence and risk of PCN in relation to serum GGT levels have not been reported in previous studies. There have been studies examining the prevalence of PCN. However, they only enrolled a subset of patients with cross-sectional imaging such as computed tomography (CT) or magnetic resonance imaging (MR) [27, 28]. Few studies have reported the prevalence and incidence of PCN in participants undergoing MR as part of preventive health checkups [16, 17]. However, associations of PCN with laboratory tests such as fasting blood glucose, liver enzymes, and GGT have not been reported. Furthermore, most studies were conducted using small cohorts of less than 3,000 participants. They showed heterogeneous results not only due to different characteristics of the population itself, but also due to different imaging modality and resolution [14, 27, 28, 29].

In this study, we focused on GGT, a well-known biomarker of oxidative stress and liver dysfunction. GGT is also associated with alcoholic liver disease and metabolic dysfunction associated fatty liver disease, which could be a potential confounding factor [30, 31]. However, in multivariate analysis, serum GGT levels were associated with the risk of PCN after adjusting for alcohol consumption and metabolic diseases such as diabetes, dyslipidemia, and obesity. These results suggest the potential of GGT as an independent biomarker of PCN.

Several biological mechanisms might explain the association between serum GGT levels and the risk of PCN. Genetic alterations (such as KRAS and GNAS) and their downstream pathways are known to be involved in the development and progression of PCN [8, 32, 33, 34]. Among them, the phosphoinositide 3-kinase and AKT (PI3K/Akt) pathway, G-protein-coupled receptor (GPCR) signaling pathway, and Wnt/β-catenin signaling pathway are key signaling molecules and pathways affected by oxidative stress [35, 36, 37]. GGT, which contributes to redox homeostasis via glutathione, could be a biomarker reflecting these pathways [38, 39]. GGT not only regulates intracellular glutathione, but also produces gamma-glutamyl peptides [40]. These peptides can trigger cell growth and differentiation through calcium-sensing receptors (CaSR) present in pancreatic ductal and acinar cells, which might also contribute to the development of PCN [41, 42]

The strength of our study was that we used data from a large-scale population with a follow-up period of 10 years. Furthermore, the NHIS database contains a wide range of data, including ICD-10 diagnostic codes, prescriptions, laboratory tests, and responses to questionnaires. As a result, we were able to identify numerous confounders, including previous pancreatitis, alcohol consumption, and physical activity. However, results of our study should be interpreted in light of the following limitations. First, some cases were excluded from the analysis due to missing data. Although the proportion of individuals with missing data in the initially screened population was relatively low (5.5%, n = 160,987), characteristics of this population might have influenced our study results. Second, although we excluded individuals who were diagnosed with PCN within 1-year lag period, reverse causality might not have been eliminated. Third, despite various covariates utilized in our analyses, residual confounding factors might be present. Fourth, we were unable to classify the type of PCN based on imaging or pathologic findings due to limited data. Finally, our findings were based on the Korean population, not other ethnicities.

In conclusion, this nationwide population study showed that higher serum GGT level was independently associated with an increased risk of PCN in a dose-response manner. Our findings suggest the potential of serum GGT level as a biomarker for early detection of PCN which might have a risk of malignancy. The development of such biomarkers will lead to earlier detection of PCN that has been detected incidentally and contribute to more established guidance on the management of PCN.

## Data Availability

All data produced in the present study are available upon reasonable request to the authors

## • Acknowledgement

None

## • Conflicts of Interest

Authors have no conflicts of interest relevant to this study to disclose.

## Notes

### Competing Interest Statement

The authors have declared no competing interest.

### Funding Statement

This study did not receive any funding

### Author Declarations

This study was approved by the Institutional Review Board (IRB) of Seoul National University Hospital (IRB No. H-2406-052-1542)

## References

1 Brugge WR, Lauwers GY, Sahani D, Fernandez-del Castillo C, Warshaw AL. Cystic neoplasms of the pancreas. N Engl J Med 2004;351:1218–26.

2 Uehara H, Nakaizumi A, Ishikawa O, Iishi H, Tatsumi K, Takakura R, et al. Development of ductal carcinoma of the pancreas during follow-up of branch duct intraductal papillary mucinous neoplasm of the pancreas. Gut 2008;57:1561–5.

3 Oyama H, Tada M, Takagi K, Tateishi K, Hamada T, Nakai Y, et al. Long-term Risk of Malignancy in Branch-Duct Intraductal Papillary Mucinous Neoplasms. Gastroenterology 2020;158:226–37.e5.

4 Kwong WT, Hunt GC, Fehmi SM, Honerkamp-Smith G, Xu R, Lawson RD, et al. Low Rates of Malignancy and Mortality in Asymptomatic Patients With Suspected Neoplastic Pancreatic Cysts Beyond 5 Years of Surveillance. Clin Gastroenterol Hepatol 2016;14:865–71.

5 Crippa S, Fernández-Del Castillo C, Salvia R, Finkelstein D, Bassi C, Domínguez I, et al. Mucin-producing neoplasms of the pancreas: an analysis of distinguishing clinical and epidemiologic characteristics. Clin Gastroenterol Hepatol 2010;8:213–9.

6 Vege SS, Ziring B, Jain R, Moayyedi P. American gastroenterological association institute guideline on the diagnosis and management of asymptomatic neoplastic pancreatic cysts. Gastroenterology 2015;148:819–22; quize12-3.

7 Tanaka M, Fernández-Del Castillo C, Kamisawa T, Jang JY, Levy P, Ohtsuka T, et al. Revisions of international consensus Fukuoka guidelines for the management of IPMN of the pancreas. Pancreatology 2017;17:738–53.

8 Noë M, Niknafs N, Fischer CG, Hackeng WM, Beleva Guthrie V, Hosoda W, et al. Genomic characterization of malignant progression in neoplastic pancreatic cysts. Nat Commun 2020;11:4085.

9 Yip-Schneider MT, Simpson R, Carr RA, Wu H, Fan H, Liu Z, et al. Circulating Leptin and Branched Chain Amino Acids-Correlation with Intraductal Papillary Mucinous Neoplasm Dysplastic Grade. J Gastrointest Surg 2019;23:966–74.

10 Ciprani D, Morales-Oyarvide V, Qadan M, Hank T, Weniger M, Harrison JM, et al. An elevated CA 19-9 is associated with invasive cancer and worse survival in IPMN. Pancreatology 2020;20:729–35.

11 Collet L, Ghurburrun E, Meyers N, Assi M, Pirlot B, Leclercq IA, et al. Kras and Lkb1 mutations synergistically induce intraductal papillary mucinous neoplasm derived from pancreatic duct cells. Gut 2020;69:704–14.

12 Gaiser RA, Pessia A, Ateeb Z, Davanian H, Fernández Moro C, Alkharaan H, et al. Integrated targeted metabolomic and lipidomic analysis: A novel approach to classifying early cystic precursors to invasive pancreatic cancer. Sci Rep 2019;9:10208.

13 European evidence-based guidelines on pancreatic cystic neoplasms. Gut 2018;67:789–804.

14 Moris M, Bridges MD, Pooley RA, Raimondo M, Woodward TA, Stauffer JA, et al. Association Between Advances in High-Resolution Cross-Section Imaging Technologies and Increase in Prevalence of Pancreatic Cysts From 2005 to 2014. Clin Gastroenterol Hepatol 2016;14:585–93.e3.

15 Zerboni G, Signoretti M, Crippa S, Falconi M, Arcidiacono PG, Capurso G. Systematic review and meta-analysis: Prevalence of incidentally detected pancreatic cystic lesions in asymptomatic individuals. Pancreatology 2019;19:2–9.

16 de Jong K, Nio CY, Hermans JJ, Dijkgraaf MG, Gouma DJ, van Eijck CH, et al. High prevalence of pancreatic cysts detected by screening magnetic resonance imaging examinations. Clin Gastroenterol Hepatol 2010;8:806–11.

17 Kromrey ML, Bülow R, Hübner J, Paperlein C, Lerch MM, Ittermann T, et al. Prospective study on the incidence, prevalence and 5-year pancreatic-related mortality of pancreatic cysts in a population-based study. Gut 2018;67:138–45.

18 Kim YG, Han K, Jeong JH, Roh SY, Choi YY, Min K, et al. Metabolic Syndrome, Gamma-Glutamyl Transferase, and Risk of Sudden Cardiac Death. J Clin Med 2022;11.

19 Baek HS, Kim B, Lee SH, Lim DJ, Kwon HS, Chang SA, et al. Long-Term Cumulative Exposure to High γ-Glutamyl Transferase Levels and the Risk of Cardiovascular Disease: A Nationwide Population-Based Cohort Study. Endocrinol Metab (Seoul) 2023;38:770–81.

20 Hong SH, Lee JS, Kim JA, Lee YB, Roh E, Yu JH, et al. Gamma-glutamyl transferase variability and the risk of hospitalisation for heart failure. Heart 2020;106:1080–6.

21 Lee CH, Han K, Kim DH, Kwak MS. Repeatedly elevated γ-glutamyltransferase levels are associated with an increased incidence of digestive cancers: A population-based cohort study. World J Gastroenterol 2021;27:176–88.

22 Liao W, Yang Y, Yang H, Qu Y, Song H, Li Q. Circulating gamma-glutamyl transpeptidase and risk of pancreatic cancer: A prospective cohort study in the UK Biobank. Cancer Med 2023;12:7877–87.

23 Song SO, Jung CH, Song YD, Park CY, Kwon HS, Cha BS, et al. Background and data configuration process of a nationwide population-based study using the korean national health insurance system. Diabetes Metab J 2014;38:395–403.

24 Cheol Seong S, Kim YY, Khang YH, Heon Park J, Kang HJ, Lee H, et al. Data Resource Profile: The National Health Information Database of the National Health Insurance Service in South Korea. Int J Epidemiol 2017;46:799–800.

25 Kang HT. Current Status of the National Health Screening Programs in South Korea. Korean J Fam Med 2022;43:168–73.

26 von Elm E, Altman DG, Egger M, Pocock SJ, Gøtzsche PC, Vandenbroucke JP. The Strengthening the Reporting of Observational Studies in Epidemiology (STROBE) Statement: guidelines for reporting observational studies. Int J Surg 2014;12:1495–9.

27 Laffan TA, Horton KM, Klein AP, Berlanstein B, Siegelman SS, Kawamoto S, et al. Prevalence of unsuspected pancreatic cysts on MDCT. AJR Am J Roentgenol 2008;191:802–7.

28 Lee KS, Sekhar A, Rofsky NM, Pedrosa I. Prevalence of incidental pancreatic cysts in the adult population on MR imaging. Am J Gastroenterol 2010;105:2079–84.

29 Zhang XM, Mitchell DG, Dohke M, Holland GA, Parker L. Pancreatic cysts: depiction on single-shot fast spin-echo MR images. Radiology 2002;223:547–53.

30 Green RM, Flamm S. AGA technical review on the evaluation of liver chemistry tests. Gastroenterology 2002;123:1367–84.

31 McCullough AJ. Update on nonalcoholic fatty liver disease. J Clin Gastroenterol 2002;34:255–62.

32 Springer S, Wang Y, Dal Molin M, Masica DL, Jiao Y, Kinde I, et al. A combination of molecular markers and clinical features improve the classification of pancreatic cysts. Gastroenterology 2015;149:1501–10.

33 Wu J, Matthaei H, Maitra A, Dal Molin M, Wood LD, Eshleman JR, et al. Recurrent GNAS mutations define an unexpected pathway for pancreatic cyst development. Sci Transl Med 2011;3:92ra66.

34 Patra KC, Kato Y, Mizukami Y, Widholz S, Boukhali M, Revenco I, et al. Mutant GNAS drives pancreatic tumourigenesis by inducing PKA-mediated SIK suppression and reprogramming lipid metabolism. Nat Cell Biol 2018;20:811–22.

35 Koundouros N, Poulogiannis G. Phosphoinositide 3-Kinase/Akt Signaling and Redox Metabolism in Cancer. Front Oncol 2018;8:160.

36 Wu D, Casey PJ. GPCR-Gα13 Involvement in Mitochondrial Function, Oxidative Stress, and Prostate Cancer. Int J Mol Sci 2024;25.

37 Chen Y, Chen M, Deng K. Blocking the Wnt/β[catenin signaling pathway to treat colorectal cancer: Strategies to improve current therapies (Review). Int J Oncol 2023;62.

38 Forman HJ, Zhang H, Rinna A. Glutathione: overview of its protective roles, measurement, and biosynthesis. Mol Aspects Med 2009;30:1–12.

39 Chai YC, Mieyal JJ. Glutathione and Glutaredoxin-Key Players in Cellular Redox Homeostasis and Signaling. Antioxidants (Basel) 2023;12.

40 Ikeda Y, Taniguchi N. Gene expression of gamma-glutamyltranspeptidase. Methods Enzymol 2005;401:408–25.

41 Rácz GZ, Kittel A, Riccardi D, Case RM, Elliott AC, Varga G. Extracellular calcium sensing receptor in human pancreatic cells. Gut 2002;51:705–11.

42 Goralski T, Ram JL. Extracellular Calcium Receptor as a Target for Glutathione and Its Derivatives. Int J Mol Sci 2022;23.

