## Supplementary material for "Serum gamma-glutamyl transferase level is associated with the risk of pancreatic cystic neoplasms: A nationwide cohort study": Table S1, Table S2, Table S3

**Supplemental Table 1.** International Classification of Diseases–10th Revision, Clinical Modification (ICD-10-CM) diagnostic codes for pancreatic diseases

|  | ICD-10-CM diagnostic codes |
| --- | --- |
| Pancreatitis | K85 Acute pancreatitis  K86.0 Alcoholic pancreatitis  K86.1 Chronic pancreatitis |
| Pseudocysts | K86.3 Pancreatic pseudocyst |
| Pancreatic cancer | C25 Malignant neoplasm of pancreas  C25.0 Malignant neoplasm of head of pancreas  C25.1 Malignant neoplasm of body of pancreas  C25.2 Malignant neoplasm of tail of pancreas  C25.3 Malignant neoplasm of pancreatic duct  C25.7 Malignant neoplasm of other parts of pancreas  C25.9 Malignant neoplasm of pancreas, unspecified |
| Pancreas cystic neoplasms | K86.2 Pancreas cyst  D13.6 Cystic neoplasm of pancreas, benign  D37.7 Cystic neoplasm of pancreas, uncertain behavior |

**Supplemental Table 2.** Definitions of comorbidities

|  | Definition |
| --- | --- |
| Diabetes mellitus | (1) ICD-10 codes E11–14 with prescriptions for anti-diabetic agents  or (2) fasting blood glucose level ≥ 126 mg/dL |
| Hypertension | (1) ICD-10 codes I10‒I13 and I15 with prescriptions for anti-hypertensive agents  or (2) systolic BP ≥ 140 mmHg or diastolic BP ≥ 90 mmHg |
| Dyslipidemia | (1) ICD-10 code E78 with prescriptions for lipid-lowering agents  or (2) total cholesterol level ≥ 240 mg/dL |
| Chronic kidney disease | (1) NHIS codes: V001 (hemodialysis), V003 (peritoneal dialysis), V005 (kidney transplantation)  or (2) eGFR of < 60 mL/min/1.73m^2^ by Modification of Diet in Renal Disease (MDRD) equation |

BP, blood pressure; CKD, chronic kidney disease; eGFR, estimated glomerular filtration rate; ICD-10, International Classification of Diseases-Tenth Revision.

**Supplemental Table 3.** Multivariate regression analysis of risk of PCN: Decile

| GGT | N | PCN | Duration, years | IR, per 1,000 PY | HR (95% CI) | | |
| --- | --- | --- | --- | --- | --- | --- | --- |
|  |  |  |  |  | Model 1 | Model 2 | Model 3 |
| D1 | 249126 | 2408 | 2487747.57 | 0.97 | 1(Ref.) | 1(Ref.) | 1(Ref.) |
| D2 | 277172 | 2859 | 2781081.13 | 1.03 | 1.061 (1.005,1.12) | 1.06 (1.004,1.119) | 1.063 (1.006,1.122) |
| D3 | 302115 | 3126 | 3035013.55 | 1.03 | 1.063 (1.008,1.121) | 1.047 (0.993,1.104) | 1.05 (0.996,1.108) |
| D4 | 205384 | 2125 | 2056766.57 | 1.03 | 1.067 (1.007,1.132) | 1.077 (1.016,1.141) | 1.079 (1.018,1.144) |
| D5 | 275236 | 3082 | 2762638.39 | 1.12 | 1.152 (1.092,1.215) | 1.125 (1.067,1.187) | 1.128 (1.069,1.191) |
| D6 | 259647 | 2769 | 2604856.26 | 1.06 | 1.098 (1.039,1.159) | 1.087 (1.029,1.148) | 1.089 (1.03,1.151) |
| D7 | 306146 | 3513 | 3073771.44 | 1.14 | 1.18 (1.12,1.243) | 1.134 (1.076,1.194) | 1.135 (1.076,1.196) |
| D8 | 258577 | 2969 | 2591226.26 | 1.15 | 1.184 (1.122,1.249) | 1.159 (1.098,1.223) | 1.158 (1.096,1.224) |
| D9 | 259796 | 2981 | 2598319.1 | 1.15 | 1.186 (1.124,1.251) | 1.167 (1.105,1.231) | 1.163 (1.1,1.229) |
| D10 | 262466 | 3108 | 2577377.24 | 1.21 | 1.251 (1.187,1.32) | 1.244 (1.18,1.312) | 1.237 (1.17,1.308) |

Model 1,	Not adjusted.

Model 2,	Adjusted for age and sex.

Model 3,	Adjusted for age, sex, lowest income quartile, BMI, smoking, drinking, regular exercise, diabetes mellitus, hypertension, dyslipidemia, and chronic kidney disease.
